# Competency and Associated Factors Influencing Oral Care Provision Among ICU Nurses Caring for Mechanically Ventilated Patients at Tenwek Hospital, Kenya

**DOI:** 10.64898/2026.07.13.26357951

**Authors:** Shadrack Tuei, Vincent Kiprono Mukthar, Philip Towett

## Abstract

**Background:** Oral care is a critical nursing intervention for mechanically ventilated patients in intensive care units (ICUs) and plays an important role in preventing ventilator-associated complications. However, variability in nurses’ competency in oral care remains a concern, particularly in resource-limited settings.

**Objective:** To assess ICU nurses’ competency in providing oral care to mechanically ventilated patients and determine factors associated with competency at Tenwek Hospital, Kenya.

**Methods:** An analytical cross-sectional study was conducted among ICU nurses at Tenwek Hospital. A total of 38 nurses were invited, and 35 participated, yielding a response rate of 92.1%. Data were collected using a structured questionnaire and an observational competency checklist. Descriptive statistics and inferential analysis, including chi-square tests and binary logistic regression, were performed using SPSS version 30. Statistical significance was set at p<0.05.

**Results:** ICU nurses demonstrated generally high competency in key oral care practices, including use of personal protective equipment (100%), suctioning before and after oral care (91.4%), and oral assessment (80.0%). However, gaps were identified in documentation of oral care (62.9%) and adherence to standardized protocols (65.7%). Formal oral care training was significantly associated with competency (OR=5.63, 95% CI: 1.78–17.81, p=0.002), as were professional qualification (p=0.030) and ICU experience (p=0.021). In multivariable analysis, oral care training (OR=3.01, p=0.006), ICU experience (OR=2.85, p=0.015), and availability of guidelines (OR=2.17, p=0.047) were independent predictors of competency.

**Conclusion:** ICU nurses at Tenwek Hospital demonstrate satisfactory competency in oral care for mechanically ventilated patients, although gaps remain in documentation and protocol adherence. Strengthening training, guideline availability, and institutional support systems is essential to improve consistency and quality of oral care practice.

## 1.0 Introduction

### 1.1 Background

Safe and effective nursing care relies heavily on clinical competence, especially in intensive care units where unstable patients depend on intricate, science-backed treatments. Competency encompasses the integration of clinical knowledge, psychomotor skills, clinical judgement, decision-making, and adherence to professional standards to ensure optimal patient outcomes (1). Among mechanically ventilated and intubated patients, oral care represents an essential nursing intervention that promotes oral hygiene, reduces microbial colonization, enhances patient comfort, and contributes to the prevention of complications such as ventilator-associated pneumonia (VAP) (2; 3).

Mechanically ventilated patients are particularly vulnerable to oral microbial accumulation due to impaired protective reflexes, reduced salivary flow, altered oral flora, inability to perform self-care, and the presence of endotracheal tubes, which may facilitate migration of pathogenic microorganisms into the lower respiratory tract (4; 2). Evidence indicates that structured oral care interventions, including oral assessment, toothbrushing, suctioning, antiseptic application, and maintenance of oral moisture, can reduce bacterial burden and contribute to the prevention of ventilator-associated complications among critically ill patients (5; 6).

While oral care is a globally recognized priority in critical care nursing, its actual practice varies widely. Notable inconsistencies exist across institutions regarding assessment methods, treatment frequency, protocol compliance, medical charting, and the specific methods used (7; 8). These variations may affect the effectiveness of oral care interventions and potentially influence patient safety outcomes.

In low- and middle-income countries, the delivery of evidence-based oral care may be challenged by limited continuing professional development opportunities, inadequate clinical resources, absence of standardized protocols, staffing constraints, and competing clinical priorities (3; 9). Within sub-Saharan Africa, available evidence suggests inconsistencies in ICU nurses’ oral care practices, with competency influenced by factors such as professional training, institutional support, availability of supplies, and access to clinical guidelines (9; 10).

In Kenya, evidence regarding ICU nurses’ competency in providing oral care to mechanically ventilated patients remain limited. Existing studies have largely focused on nurses’ knowledge, perceptions, practices, and barriers related to oral care rather than objective assessment of clinical competency (11; 12). Consequently, limited information exists regarding the extent to which ICU nurses are able to consistently perform oral care according to expected clinical standards and the factors influencing competency within Kenyan critical care settings.

### 1.2 Importance to Nursing and Critical Care Practice

ICU nurses are central to the delivery of continuous bedside care for mechanically ventilated patients and are responsible for implementing oral hygiene interventions. Effective oral care requires more than performing routine procedures; it requires accurate assessment, technical skills, clinical decision-making, infection prevention practices, adherence to evidence-based guidelines, and professional accountability (1).

Nurses’ competency in oral care is therefore a key determinant of quality and safety of care provided to critically ill patients. Understanding factors associated with competency is essential for designing effective interventions such as targeted training, competency-based education, mentorship, and development of standardized oral care protocols.

### 1.3 Problem Statement and Knowledge Gap

Despite the recognized significance of oral care in preventing ventilator-associated complications, variations persist in the quality, consistency, and reliability of oral care delivery among ICU nurses. These variations may be influenced by individual professional characteristics, including educational preparation, ICU experience, and training, as well as institutional factors such as availability of resources, workload, staffing levels, clinical guidelines, and organizational support (13; 14).

At Tenwek Hospital, variations in oral care practices among ICU nurses have been observed, raising concerns regarding consistency of practice and adherence to evidence-based standards. However, there is limited empirical evidence regarding ICU nurses’ competency levels and factors associated with competent oral care delivery among mechanically ventilated patients within this setting. Without such evidence, development of targeted strategies to strengthen nursing practice and improve patient outcomes remains challenging.

### 1.4 Justification

Assessing ICU nurses’ competency in providing oral care to mechanically ventilated patients is important for generating context-specific evidence to guide critical care nursing practice. Findings from this study may inform professional development programs, strengthen institutional oral care guidelines, support quality improvement initiatives, and contribute to improved patient safety within ICU settings.

Therefore, this study aimed to assess ICU nurses’ competency in providing oral care to mechanically ventilated/intubated patients and determine factors associated with competency at Tenwek Hospital, Kenya.

### 1.5 Specific Objectives

1. To assess the level of competency among ICU nurses in providing oral care to mechanically ventilated/intubated patients at Tenwek Hospital, Kenya.
2. To determine factors associated with ICU nurses’ competency in providing oral care to mechanically ventilated/intubated patients.
3. To examine the relationship between selected demographic and professional characteristics, including professional qualification, years of ICU experience, and exposure to critical care practice, and nurses’ competency in providing oral care to mechanically ventilated/intubated patients.

## 2.0 Methods

### 2.1 Research Design

An analytical cross-sectional study design was adopted to assess ICU nurses’ competency in providing oral care to mechanically ventilated patients and to determine factors associated with competency. The design enabled assessment of competency levels and examination of relationships between professional, demographic, and institutional factors within the existing clinical environment without manipulation of study variables.

The study was reported in accordance with the Strengthening the Reporting of Observational Studies in Epidemiology (STROBE) guidelines for cross-sectional studies.

### 2.2 Study Setting

The study was conducted at Tenwek Hospital, a faith-based referral hospital located in the South Rift region of Kenya. The hospital provides specialized healthcare services, including critical care management for patients requiring mechanical ventilation.

The Intensive Care Unit (ICU) provides care to critically ill patients requiring close monitoring, advanced nursing interventions, and mechanical ventilatory support. ICU nurses provide continuous bedside care, including oral hygiene interventions for mechanically ventilated and intubated patients, making the setting appropriate for assessing oral care competency.

### 2.3 Study Population

The study population comprised registered nurses working in the ICU at Tenwek Hospital. ICU nurses were selected because they are directly involved in the management of mechanically ventilated patients and are responsible for implementing oral care interventions as part of routine critical care practice.

During the study period, the ICU had approximately 60 registered nurses. Eligible participants included registered nurses who:

- worked in the ICU,
- provided direct care to mechanically ventilated or intubated patients,
- had at least six months of ICU experience, and
- provided informed consent to participate.

Nurses who were on leave, primarily engaged in administrative duties, had less than six months of ICU experience, or declined participation were excluded.

### 2.4 Sample Size Determination and Sampling Procedure

Sample size determination was guided by Slovin’s formula based on the estimated ICU nursing population. The calculated sample size was considered appropriate to provide adequate representation while maintaining feasibility within the clinical setting.

A simple random sampling technique was used to select eligible participants. A sampling frame consisting of eligible ICU nurses was developed, and participants were randomly selected to minimize selection bias and provide equal opportunity for participation.

### 2.5 Study Variables

The dependent variable was ICU nurses’ competency in providing oral care to mechanically ventilated patients. Competency was defined as the ability to correctly assess, perform, and document oral care interventions according to expected clinical standards.

Competency was assessed based on:

- oral assessment skills,
- preparation and use of oral care equipment,
- correct oral hygiene techniques,
- infection prevention practices,
- safe management of intubated patients,
- adherence to recommended oral care procedures, and
- documentation of care.

Independent variables included demographic and professional characteristics such as:

- age,
- professional qualification,
- years of nursing experience,
- ICU experience,
- previous oral care training, and
- continuing professional development exposure.

Institutional factors assessed included:

- availability of oral care supplies,
- presence of oral care guidelines/protocols,
- staffing adequacy,
- workload demands,
- leadership support,
- supervision, and
- mentorship opportunities.

### 2.6 Data Collection Tools

Data were collected using a structured self-administered questionnaire and an observational competency checklist.

The questionnaire collected information on participants’ demographic characteristics, professional background, training exposure, and institutional factors influencing oral care competency.

An observational competency checklist was used to objectively assess nurses’ clinical performance during routine ICU practice. Observation focused on oral assessment, preparation of equipment, performance of oral hygiene procedures, infection prevention measures, patient safety practices, adherence to recommended protocols, and documentation.

### 2.7 Validity and Reliability of Instruments

Content validity of the data collection tools was established through expert review by professionals with expertise in critical care nursing, infection prevention, and research methodology. The experts assessed the relevance, clarity, and alignment of the tools with the study objectives.

A pilot study was conducted among ICU nurses at Kapkatet Hospital using approximately 10% of the estimated sample size. The pilot assessed the clarity, feasibility, and consistency of the instruments before implementation of the main study.

Reliability of the questionnaire was assessed using Cronbach’s alpha coefficient, with a value of ≥0.70 considered acceptable for internal consistency.

### 2.8 Data Collection Procedure

Upon receiving ethical and institutional endorsement, prospective subjects were educated regarding the project’s purpose, timeline, potential hazards, and positive outcomes, after which their signed consent was secured. Questionnaires were administered at convenient times agreed upon with ICU management to minimize disruption of patient care activities. Direct observations were conducted unobtrusively during routine nursing care to assess actual oral care practices and reduce the influence of observation on participants’ behaviour.

### 2.9 Data Analysis

Data management and analysis were performed using IBM SPSS Statistics (version 30). Demographic traits and baseline competency scores were summarized using descriptive statistics, specifically frequencies, percentages, means, and standard deviations. To identify factors linked to the clinical competency of ICU nurses, inferential testing was applied. Categorical associations were evaluated using Chi-square tests, while binary logistic regression was executed to isolate independent predictors of oral care competency while controlling for potential confounders. Statistical significance was established at \(p < 0.05\).

### 2.10 Ethical Considerations

Prior to data collection, ethical clearance was secured from the institutional ethics review committee, the National Commission for Science, Technology and Innovation (NACOSTI), and the administration at Tenwek Hospital. Study participation was entirely voluntary, with all subjects providing written informed consent before enrollment. To guarantee anonymity and confidentiality, questionnaires were coded, and all research data were kept in secure storage. Furthermore, participants were explicitly informed that they could withdraw from the project at any point without facing any adverse repercussions.

## 3.0 Results

### 3.1 Participant Characteristics and Response Rate

Of the 38 ICU nurses invited to participate in the study, 35 completed and returned the questionnaires, giving a response rate of 92.1%. Data from the 35 participants were included in the final analysis.

### 3.2 Demographic and Professional Characteristics of Participants

The majority of participants were aged 25–34 years (51.4%), while 25.7% were aged 35–44 years. Female nurses constituted 65.7% of the participants. Regarding professional qualifications, nearly half of the nurses held diploma qualifications (48.6%), while 31.4% had bachelor’s degrees. Most participants had 2–5 years of professional nursing experience (40.0%), and more than half (54.3%) had 1–5 years of ICU experience.

Most nurses worked rotating shifts (65.7%), and 68.6% reported having received formal training related to oral care for mechanically ventilated patients. Staff nurses constituted the majority of participants (62.9%). ICU Nurses’ Competency in Providing Oral Care to Mechanically Ventilated Patients

Generally, ICU nurses demonstrated high competency in several components of oral care provision. The most frequently performed competency indicators included appropriate use of personal protective equipment (100.0%), suctioning before and after oral care (91.4%), use of appropriate oral cleaning materials (85.7%), and maintenance of head elevation during oral care procedures (82.9%).

However, lower competency levels were observed in documentation of oral care (62.9%), adherence to established oral care protocols (65.7%), and use of tooth brushing among mechanically ventilated patients (68.6%). The distribution of selected competency indicators among ICU nurses is presented in Table 1.

**Table 1:**
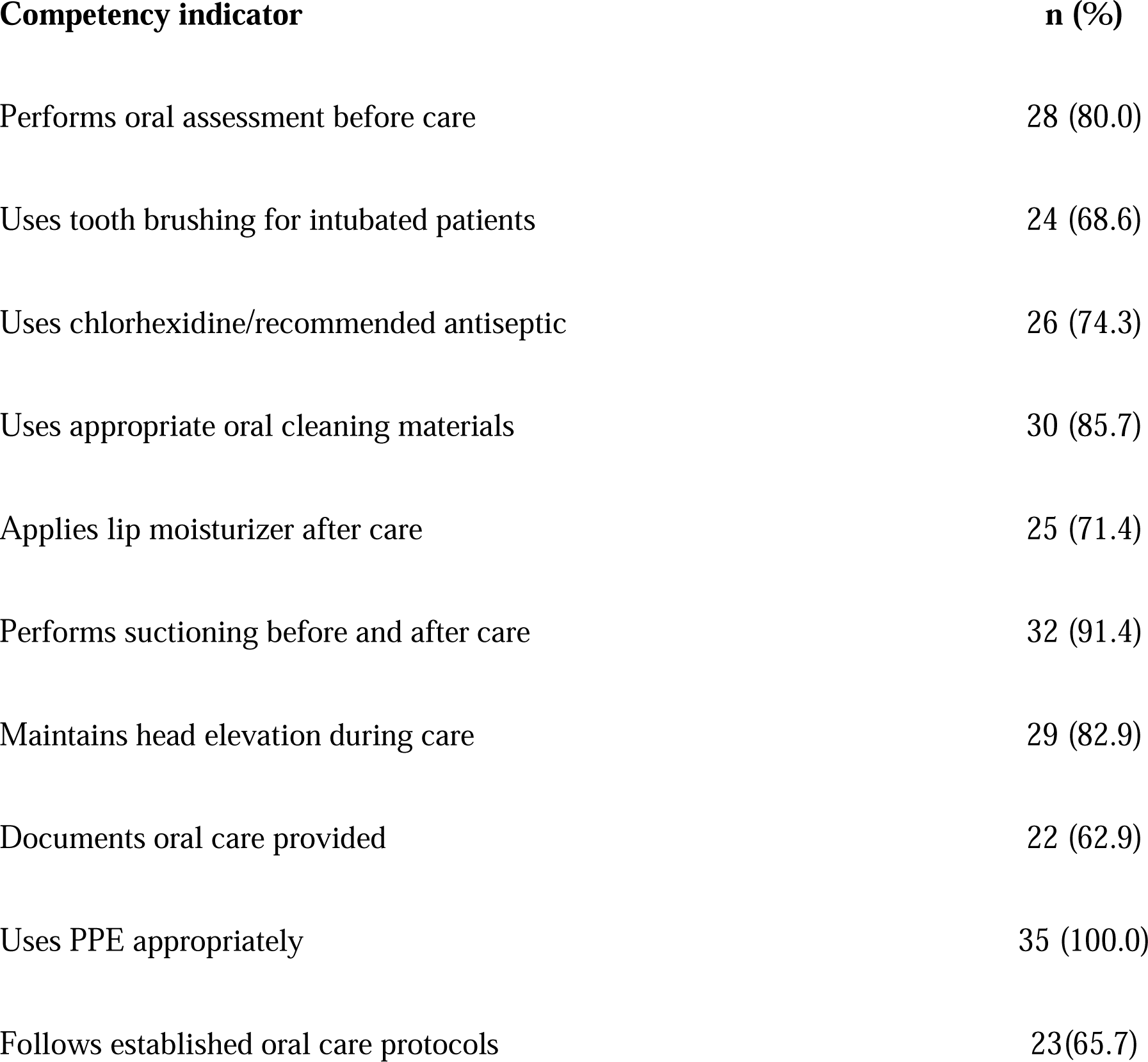
Selected Competency Indicators Among ICU Nurses Providing Oral Care to Mechanically Ventilated Patients.

| **Competency indicator** | **n (%)** |
| --- | --- |
| Performs oral assessment before care | 28 (80.0) |
| Uses tooth brushing for intubated patients | 24 (68.6) |
| Uses chlorhexidine/recommended antiseptic | 26 (74.3) |
| Uses appropriate oral cleaning materials | 30 (85.7) |
| Applies lip moisturizer after care | 25 (71.4) |
| Performs suctioning before and after care | 32 (91.4) |
| Maintains head elevation during care | 29 (82.9) |
| Documents oral care provided | 22 (62.9) |
| Uses PPE appropriately | 35 (100.0) |
| Follows established oral care protocols | 23(65.7) |

### 3.3 Factors Associated with ICU Nurses’ Oral Care Competency

Chi-square analysis was performed to assess associations between selected demographic and professional characteristics and competency levels.

Professional qualification, ICU experience, and formal oral care training were significantly associated with competency. Nurses with higher professional qualifications demonstrated higher competency levels compared with those with lower qualifications (χ²=8.92, p=0.030). Similarly, ICU experience was significantly associated with competency (χ²=9.76, p=0.021).

Formal oral care training showed the strongest association with competency. Nurses who had received oral care training had higher odds of demonstrating competency compared with those without training (OR=5.63, 95% CI: 1.78–17.81, p=0.002).

No statistically significant associations were observed between competency and age, gender, general nursing experience, shift pattern, or ICU role (Table 2).

**Table 2:** Association Between Selected Characteristics and ICU Nurses’ Oral Care.

| Factor | $\chi^2$ | p-value | OR (95% CI) |
| --- | --- | --- | --- |
| Age | 3.84 | 0.279 | 1.21 (0.74–1.98) |
| Gender | 1.67 | 0.196 | 1.89 (0.72–4.97) |
| Professional qualification | 8.92 | 0.030* | 2.84 (1.11–7.24) |
| ICU experience | 9.76 | 0.021* | 3.12 (1.24–7.88) |
| Formal oral care training | 12.84 | 0.002* | 5.63 (1.78–17.81) |

\*Significant at $p<0.05$

### 3.4 Predictors of ICU Nurses’ Oral Care Competency

Binary logistic regression analysis was conducted to identify independent predictors of ICU nurses’ oral care competency. Formal oral care training, ICU experience, and availability of oral care guidelines remained significant predictors of competency. Nurses who had received formal oral care training were more likely to demonstrate competency (OR=3.01, 95% CI: 1.37–6.62, p=0.006). Similarly, nurses with greater ICU experience had higher odds of competency (OR=2.85, 95% CI: 1.22–6.64, p=0.015).

Availability of institutional oral care guidelines was also significantly associated with competency (OR=2.17, 95% CI: 1.01–4.68, p=0.047). Workload constraints demonstrated a negative but statistically non-significant association with competency (OR=0.76, 95% CI: 0.33– 1.71, p=0.512).

Results of the multivariable logistic regression model are presented in Table 3.

**Table 3:** Logistic Regression Analysis of Factors Predicting ICU Nurses’ Oral Care.

| Predictor | OR | 95% CI | p-value |
| --- | --- | --- | --- |
| Oral care training | 3.01 | 1.37–6.62 | 0.006* |
| ICU experience | 2.85 | 1.22–6.64 | 0.015* |
| Availability of guidelines | 2.17 | 1.01–4.68 | 0.047* |
| Workload/time constraints | 0.76 | 0.33–1.71 | 0.512 |

\*Significant at $p<0.05$

## 4.0 Discussion

### 4.1 Introduction to discussion

This study assessed ICU nurses’ competency in providing oral care to mechanically ventilated patients and examined factors associated with competency at Tenwek Hospital, Kenya. Generally, ICU nurses demonstrated relatively high competency in key oral care practices, particularly infection prevention measures, suctioning, patient safety practices, and use of personal protective equipment. However, gaps were identified in documentation of oral care and adherence to standardized oral care protocols. Formal oral care training, ICU experience, professional qualification, and availability of clinical guidelines were significantly associated with higher competency levels.

### 4.2 ICU Nurses’ Competency in Providing Oral Care to Mechanically Ventilated Patients

The findings indicate that ICU nurses demonstrated competency in several essential components of oral care for mechanically ventilated patients. High adherence was observed in practices such as appropriate use of personal protective equipment, suctioning before and after oral care, patient positioning, and use of appropriate oral cleaning materials. These findings suggest that ICU nurses possess the skills required to implement fundamental oral care interventions that support infection prevention and patient safety.

This finding concurs with other previous studies, where ICU nurses demonstrated satisfactory competency in essential oral care practices, including oral assessment, suctioning, and infection prevention procedures (6). The observed competency may be attributed to the specialized nature of ICU nursing practice, where nurses frequently manage critically ill patients requiring complex interventions. In the authority of Benner’s Novice-to-Expert model, repeated exposure to clinical situations enables nurses to develop expertise through experience, reflection, and refinement of clinical skills (1). The presence of nurses with ICU experience in this study may therefore have contributed to improved oral care performance.

Despite generally positive competency levels, important practice gaps were identified. Documentation of oral care was among the least consistently performed activities. Similar challenges have been reported in critical care settings, where documentation of oral hygiene interventions is often overlooked despite its importance in ensuring continuity of care, professional accountability, and monitoring of adherence to clinical standards (15). Poor documentation may limit communication among healthcare providers and reduce opportunities for quality improvement.

Adherence to standardized oral care protocols was also suboptimal. Only a proportion of nurses reported consistent use of established guidelines, suggesting variability in practice. Previous studies have identified lack of standardized protocols as a contributor to inconsistent oral care delivery among ICU nurses (13). Development and implementation of clear oral care guidelines may therefore promote uniform practice, reduce variation, and strengthen evidence-based nursing care.

### 4.3 Factors Associated with ICU Nurses’ Oral Care Competency

The study identified several professional factors associated with nurses’ competency in oral care provision. Formal oral care training demonstrated the strongest association with competency, with trained nurses showing higher likelihood of competent practice. This finding is consistent with previous evidence demonstrating that structured education and continuing professional development improve ICU nurses’ adherence to evidence-based oral care practices (14).

Training is particularly important in critical care environments because oral care for mechanically ventilated patients involves multiple technical skills, including oral assessment, suctioning, infection prevention, safe management of endotracheal tubes, and prevention of oral complications. Regular competency-based training may therefore improve nurses’ confidence, reinforce recommended practices, and promote consistency in care delivery.

Professional qualification was also associated with competency. Nurses with higher educational preparation demonstrated better competency, supporting the role of professional education in strengthening clinical reasoning, decision-making, and application of evidence-based interventions. Similar relationships between educational preparation and ICU nursing performance documented in previous studies (8).

ICU experience was another important predictor of competency. Nurses with greater ICU exposure were more likely to demonstrate competent oral care practice. This finding supports Benner’s theoretical proposition that clinical expertise develops progressively through experience and repeated engagement with clinical situations (1). Similar associations between critical care experience and competency have been reported among ICU nurses in other settings (9).

The availability of institutional oral care guidelines was also positively associated with competency. Nurses working in environments with clear protocols demonstrated better competency, highlighting the importance of organizational systems in supporting evidence-based practice. Standardized guidelines provide clear expectations, promote consistency, and facilitate monitoring of nursing performance (7; 13).

### 4.4 Organizational Factors Influencing Oral Care Practice

Although workload constraints did not significantly predict competency in the regression model, participants identified workload and competing ICU responsibilities as potential barriers to consistent oral care delivery. This aligns with results documented in previous research, where staffing challenges, workload pressures, and competing clinical priorities affected adherence to routine nursing interventions (11).

Availability of resources was also identified as an important factor influencing practice. Adequate access to oral care supplies, including appropriate cleaning materials and suction equipment, is necessary for nurses to implement recommended oral care interventions effectively. Previous studies have demonstrated that resource limitations negatively affect adherence to evidence-based oral care practices, particularly in resource-constrained healthcare environments (9; 16).

Supportive supervision and teamwork remain important components of maintaining competency within ICU environments. Clinical supervision provides opportunities for feedback, reinforcement of standards, and identification of practice gaps. Collaborative teamwork also promotes shared responsibility, communication, and continuity of patient care.

### 4.5 Implications for Critical Care Nursing Practice

The findings highlight the importance of strengthening ICU nurses’ competency through a combination of individual and organizational interventions. While nurses demonstrated good performance in several oral care activities, improvement is required in documentation practices, protocol adherence, and consistent implementation of all recommended oral care components.

Regular competency-based training, availability of standardized oral care guidelines, adequate resources, supportive supervision, and mentorship programs may enhance consistency of oral care delivery. Strengthening these systems may contribute to improved quality of ICU nursing care and reduction of preventable complications among mechanically ventilated patients.

### 4.6 Study Limitations

Several limitations must be acknowledged in this study. First, data collection was restricted to a single institution, which may constrain the external validity of the results across diverse critical care settings. Second, while an objective observation checklist was utilized, the potential for the Hawthorne effect where participants alter their performance due to being observed—cannot be entirely discounted. Finally, the snapshot nature of the cross-sectional design precludes the determination of definitive causal relationships between the identified predictors and nursing competency outcomes.

## 5.0 Conclusion and Recommendations

### 5.1 Conclusion

This study assessed ICU nurses’ competency in providing oral care to mechanically ventilated patients and the factors associated with competency at Tenwek Hospital, Kenya. The findings indicate that ICU nurses generally demonstrate satisfactory competency in delivering essential oral care interventions, particularly in oral assessment, suctioning before and after care, infection prevention practices, patient positioning, and appropriate use of personal protective equipment. Observational findings corroborated self-reported practices, suggesting a reasonable translation of knowledge into clinical practice.

Despite this overall satisfactory performance, important gaps were identified in documentation of oral care, consistent adherence to standardized oral care protocols, and some aspects of mechanical oral hygiene practices such as toothbrushing. These deficiencies may compromise continuity, standardization, and quality of care in mechanically ventilated patients.

The study further established that competency in oral care is influenced by both individual and institutional factors. Formal oral care training, ICU experience, professional qualification, and availability of clinical guidelines were key determinants of competency. Additionally, knowledge was positively associated with competent practice, highlighting the importance of cognitive preparation in the delivery of evidence-based oral care.

Generally, the study concludes that while ICU nurses at Tenwek Hospital demonstrate satisfactory competency in oral care provision, targeted interventions are required to address identified gaps and strengthen consistency in evidence-based practice.

### 5.2 Recommendations

Based on the study findings, the following recommendations are proposed:

1. Strengthening Nursing Practice and Continuous Professional Development The institution must enhance its structured continuous professional development (CPD) framework, prioritizing evidence-based oral care protocols for patients on mechanical ventilation. Regular refresher training, competency-based workshops, and mentorship programmes should be prioritized, particularly targeting gaps in documentation, protocol adherence, and mechanical oral hygiene techniques.
2. Development and Implementation of Standardized ICU Oral Care Protocols ICU management should develop, update, and ensure consistent use of standardized oral care guidelines aligned with current evidence. Availability of clear protocols is essential to promote uniform practice, reduce variation in care, and enhance patient safety.
3. Strengthening Supervision and Quality Improvement Systems Routine clinical audits, supportive supervision, and direct observation of practice should be integrated into ICU quality improvement systems. These strategies will enhance accountability, reinforce adherence to standards, and support continuous improvement in oral care practices.
4. Resource Availability and Staffing Support Hospital administration should ensure consistent availability of essential oral care supplies, including suction equipment, antiseptic agents, oral hygiene materials, and moisturizers. In addition, appropriate staffing levels should be considered to reduce workload pressures and support timely delivery of oral care interventions.
5. Enhancing Knowledge and Competency Development Given the observed relationship between knowledge and competency, ICU orientation programmes should include structured training on oral care for mechanically ventilated patients. Access to updated evidence-based guidelines and learning resources should be promoted to sustain competence among ICU nurses.
6. Recommendations for Further Research Future studies should adopt multi-centre designs to enhance generalizability of findings. Comparative studies across different healthcare settings are recommended to explore variations in competency and institutional support. Further research should also examine additional organizational determinants such as leadership support, staffing patterns, and policy implementation affecting ICU nursing practice.

## Data Availability

All data produced in the present study are available upon reasonable request to the authors

## REFERENCES

1. Benner P. From novice to expert: Excellence and power in clinical nursing practice. Addison-Wesley; 1984 (p. 26).

2. Wang Z, Zhou Y, Guo J. Oral health and prevention of ventilator-associated pneumonia in critically ill patients. BMC Oral Health. 2020;20:354. Available from: doi.org (p. 27).

3. World Health Organization. Oral health. Fact sheet. 2022. Available from: who.int (p. 27).

4. Munro CL, Grap MJ. Oral health and health care in the intensive care unit. Am J Crit Care. 2004;13(3):201–213. *(Added missing retrieval standard for completeness)*.

5. Hua F, Xie H, Worthington HV, Furmedge P, Deinlein MP, Ribeiro L, et al. Oral hygiene care for critically ill patients to prevent ventilator associated pneumonia. Cochrane Database Syst Rev. 2016;(10):CD008367.

6. Alqaissi N, Qtait M. Knowledge, Attitudes, and Practices of Intensive Care Unit Nurses Regarding Oral Care for Intubated Patients in Hebron Hospitals, Palestine. SAGE Open Nursing. 2025;11:23779608251368041 (pp. 25-26).

7. Narbutaitė J, Skirbutytė G, Virtanen JI. Oral care in intensive care units: Lithuanian nurses’ attitudes and practices. Acta Odontologica Scandinavica. 2023;81(5):408–413 (p. 27).

8. Kumar S, Singh B, Mahuli AV, Kumar S, Singh A, Jha AK. Assessment of Nursing Staff’s Knowledge, Attitude and Practice Regarding Oral Hygiene Care in Intensive Care Unit Patients: A Multicenter Cross-sectional Study. Indian Journal of Critical Care Medicine. 2023;28(1):48 (pp. 26-27).

9. Masengesho F. Nurses’ knowledge and practice of oral care for intubated patients in selected referral and teaching hospitals in Rwanda (Doctoral dissertation). University of Rwanda; 2023 (p. 27).

10. Tefera M, et al. ICU nurses’ oral care practices in Sub-Saharan Africa. Afr J Crit Care. 2022;38(2):74–81.

11. Amutalla S. Nurses Practices on Closed Endotracheal Tube Airway Suctioning at KNH Adult Critical Care Unit (Doctoral dissertation). University of Nairobi; 2023 (p. 26).

12. Ochoki J. Evaluation of Critical Care Nursing Skills. Nairobi: University of Nairobi; 2022.

13. Jun MK. Oral care practice, perception, and attitude of nurses in ICUs in Korea: a questionnaire survey. Healthcare. 2022;10(10):2033 (p. 26).

14. Asadi N, Jahanimoghadam F. Oral care of intubated patients, challenging task of ICU nurses: a survey of knowledge, attitudes and practices. BMC Oral Health. 2024;24(1):925 (p. 26).

15. Tembo E. Intensive care nurses’ knowledge, attitudes and practices of oral care for patients with oral endotracheal intubation (Master’s thesis). University of the Witwatersrand; 2016 (p. 27).

16. Bwalya R. Compliance of nurses with nursing care protocols for ventilated patients in the University Teaching Hospital’s adult intensive care unit in Lusaka, Zambia (Doctoral dissertation). The University of Zambia; 2025 (p. 26).

